# Artificial Intelligence-Enabled Echocardiographic Assessment of Right Ventricular Function

**DOI:** 10.64898/2026.01.20.26344425

**Authors:** Márton Tokodi, Bryan He, Ádám Szijártó, Andrea Ferencz, Kai Shiida, Máté Tolvaj, Alexandra Fábián, Milos Vukadinovic, Béla Merkely, Susan Cheng, Chung-Lieh Hung, Attila Kovács, David Ouyang

## Abstract

**Background:** Right ventricular (RV) function is an important predictor of morbidity and mortality in various cardiovascular conditions. Nevertheless, its echocardiographic assessment is challenging due to its complex anatomy and location in the chest, resulting in limited inter-observer reproducibility.

**Objectives:** We aimed to develop a novel deep learning model – **EchoNet-RV** – to segment the RV in apical 4-chamber view (A4C) echocardiographic videos and estimate RV fractional area change (RVFAC).

**Methods:** For training EchoNet-RV, 7,169 expert-annotated A4C echocardiographic videos were used. The model’s performance was evaluated on a held-out internal test set of 1,320 A4C videos and two international external test sets of 3,107 and 1,077 A4C videos from two separate centers. Additionally, the associations between the predicted RVFAC values and the composite endpoint of heart failure hospitalization or all-cause death were also analyzed in the first external test set.

**Results:** EchoNet-RV segmented the RV with Dice coefficients of 0.893 (0.891–0.895), 0.797 (0.796–0.798), and 0.788 (0.785–0.790) and predicted RVFAC with mean absolute errors of 5.795 (5.560–6.031), 5.830 (5.692–5.970), and 6.362 (6.064–6.660) percentage points in the held-out test set and the two external test sets, respectively. In 500 randomly selected videos from the external test sets, EchoNet-RV’s prediction error was significantly lower than the inter-observer variability (p<0.001). Moreover, it identified RVFAC <35% with areas under the receiver operating characteristic curve of 0.859 (0.843–0.876), 0.725 (0.710–0.740), and 0.684 (0.653–0.713) in the three test sets. EchoNet-RV also outperformed two multi-task models, EchoPrime and PanEcho, in estimating RVFAC and identifying RV dysfunction in the external test sets. In the first external test set, predicted RVFAC values were inversely associated with the composite endpoint (adjusted HR: 0.948 [0.917–0.979], p<0.001), independent of age, sex, cardiovascular risk factors, and left ventricular systolic function.

**Conclusions:** EchoNet-RV enables the rapid and automated assessment of RVFAC, with strong potential to become a valuable tool for the echocardiographic evaluation of RV function and disease surveillance.

**CONDENSED ABSTRACT:** In this study, we developed EchoNet-RV, an echocardiography-based DL model for automated RV segmentation and RVFAC estimation, and evaluated its performance on two international external datasets. EchoNet-RV demonstrated robust performance in RV segmentation, RVFAC estimation, and RV dysfunction detection, with prediction errors significantly lower than inter-observer variability. It also outperformed two multi-task models, EchoPrime and PanEcho, in estimating RVFAC and identifying RV dysfunction. Moreover, the model’s predictions were also associated with adverse clinical outcomes. EchoNet-RV enables rapid and automated RVFAC assessment, with strong potential to become a valuable tool for the echocardiographic evaluation of RV function and disease surveillance.

## INTRODUCTION

Right ventricular (RV) function is a key determinant of quality of life, exercise capacity, morbidity, and mortality across a wide range of cardiopulmonary diseases (1,2). Therefore, accurate detection and continuous surveillance of RV dysfunction are pivotal in routine clinical practice. To date, echocardiography remains the first-line imaging modality for assessing RV structure and function (1). However, large national and international echocardiography surveys have consistently reported that the majority of clinicians assess RV function only qualitatively (3–5). This reliance on qualitative evaluation reflects the inherent challenges of evaluating the complex anatomy and contraction pattern of the RV using two-dimensional echocardiography (6), as well as the persistent issues with standardization in image acquisition, analysis, and reporting (7). These limitations contribute to the underutilization of RV functional parameters in clinical decision-making and their limited incorporation into clinical guidelines (7).

Unlike tricuspid annular plane systolic excursion (TAPSE) and tricuspid annular peak systolic velocity by tissue Doppler imaging (S’) (i.e., the most widely used quantitative metrics), RV fractional area change (RVFAC) not only the longitudinal shortening of the RV but also its radial contractions and the contribution of the interventricular septum, resulting in a strong correlation with cardiac magnetic resonance imaging-derived RV ejection fraction (8). Accordingly, the 2025 American Society of Echocardiography guidelines for right heart assessment recommend RVFAC to be incorporated as a standard component into the echocardiographic evaluation of RV function (9). However, even in well-controlled research settings, RVFAC measurements show considerable heterogeneity and inter-observer variability (10–12). This variability arises from reliance on reader experience, inconsistencies in image quality, and persistent standardization challenges related to RVFAC measurement, as outlined above.

With the growing adoption of artificial intelligence (AI)-based semantic segmentation models in cardiovascular imaging (13–15), these technologies may offer a promising solution to current challenges by improving measurement consistency and enhancing generalizability, while also expanding access to automated RV function quantification. AI-based measurements have been shown to reduce intra-observer variability in a blinded comparison with echocardiographer assessments (16), and numerous deep learning (DL) models have been developed to automate labor-intensive manual tasks such as contouring and annotation of echocardiographic images (17–19). Beyond automation, AI-augmented qualitative and quantitative assessment of cardiac structure and function may also reveal subclinical changes in cardiac physiology (20,21).

Accordingly, we aimed to develop a novel DL model – EchoNet-RV – to segment the RV in apical 4-chamber view (A4C) echocardiographic videos and predict RVFAC (Central Illustration). We hypothesized that this model would provide reliable and reproducible RVFAC measurements in a fully automated manner, facilitating scalable and objective RV function assessment. Beyond extensively evaluating EchoNet-RV’s performance in three geographically distinct cohorts, we also aimed to analyze the associations between the predicted RVFAC values and clinical outcomes to prove the prognostic capabilities of the model.

**Central Illustration.**
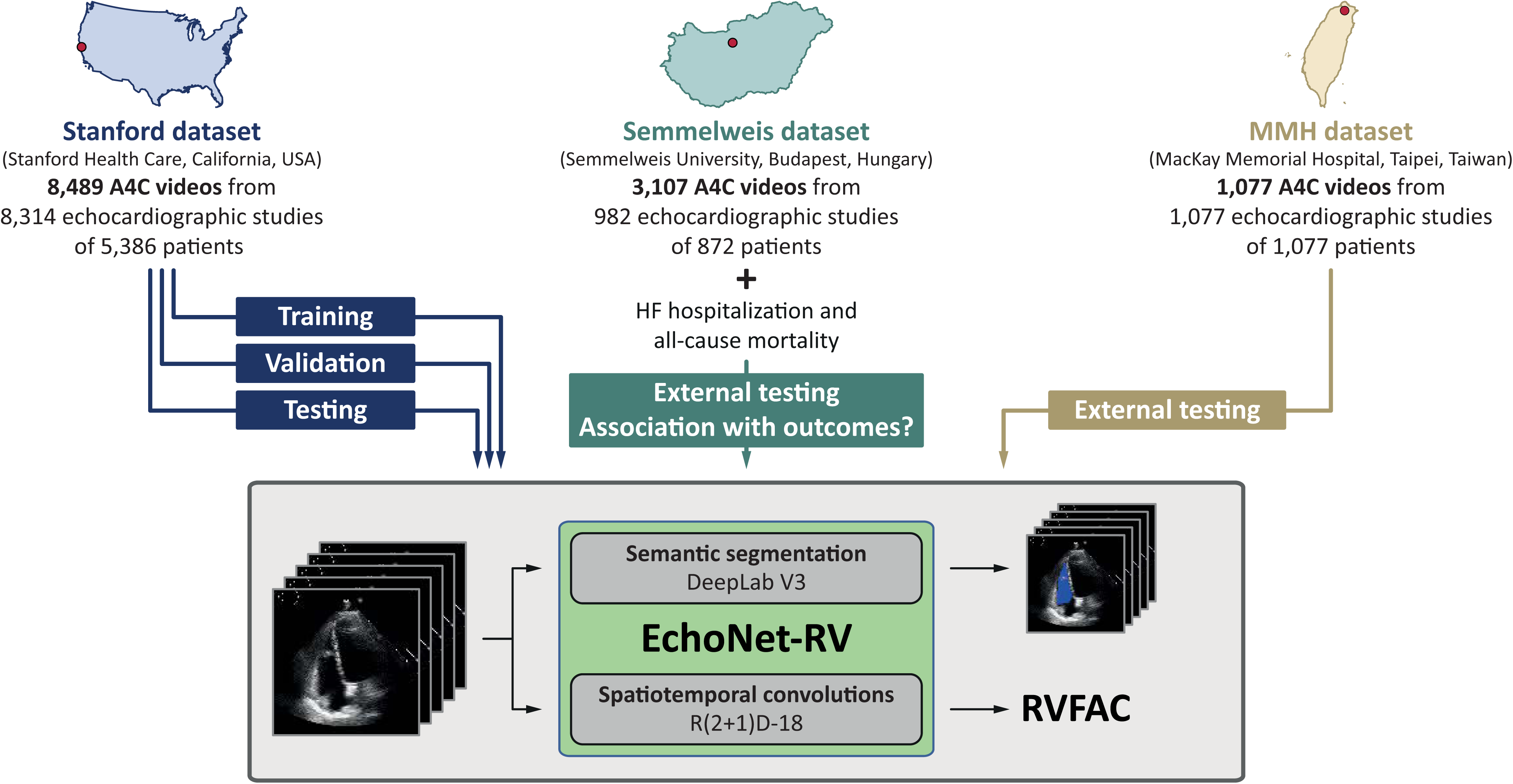
AI-enabled echocardiographic assessment of RV function. In this study, we developed a novel deep learning model – EchoNet-RV – to segment the RV in A4C echocardiographic videos and estimate RVFAC. The model was trained and internally tested in 8,489 A4C echocardiographic videos from a single healthcare system. Its performance was evaluated on two international external test sets of 3,107 and 1,077 A4C videos from two separate centers. Additionally, the associations between the predicted RVFAC values and the composite endpoint of HF hospitalization or all-cause death were also analyzed in the first external test set. A4C – apical 4-chamber view, HF – heart failure, MMH – MacKay Memorial Hospital, RV – right ventricle, RVFAC – right ventricular fractional area change

## METHODS

### Datasets

#### Stanford dataset

The Stanford dataset, used for model development and internal validation, comprised a total of 8,489 A4C videos from 8,314 studies of 5,386 patients who underwent transthoracic echocardiography between 2016 and 2018 as part of clinical care at Stanford Health Care (California, USA). Videos were acquired using Philips iE33, Philips EPIQ 5G, Philips EPIQ 7C, or Siemens Acuson SC2000 ultrasound machines. In this dataset, each video represents a unique study. Videos were randomly split into three sets of 5,892, 1,277, and 1,320 videos for training, validation, and testing, respectively. Splitting was performed at the patient level to avoid including videos from the same patient in more than one of the three sets. Clinical characteristics of the entire Stanford dataset and its three subsets are summarized in Supplemental Table 1, whereas technical details of the videos are presented in Supplemental Table 2.

#### Semmelweis dataset

The Semmelweis dataset, used for external validation and for assessing the prognostic value of the predicted RVFAC values, consisted of a total of 3,107 A4C videos from 982 studies of 872 patients who underwent transthoracic echocardiography between 2013 and 2021 at the Heart and Vascular of Semmelweis University (Budapest, Hungary). Clinical characteristics of these patients are summarized in Table 1, whereas technical details of the videos are presented in Supplemental Table 3. For patients included in the present study, mortality data were obtained from Hungary’s National Health Insurance Database, whereas information on heart failure hospitalizations was collected through chart review.

**Table 1.**
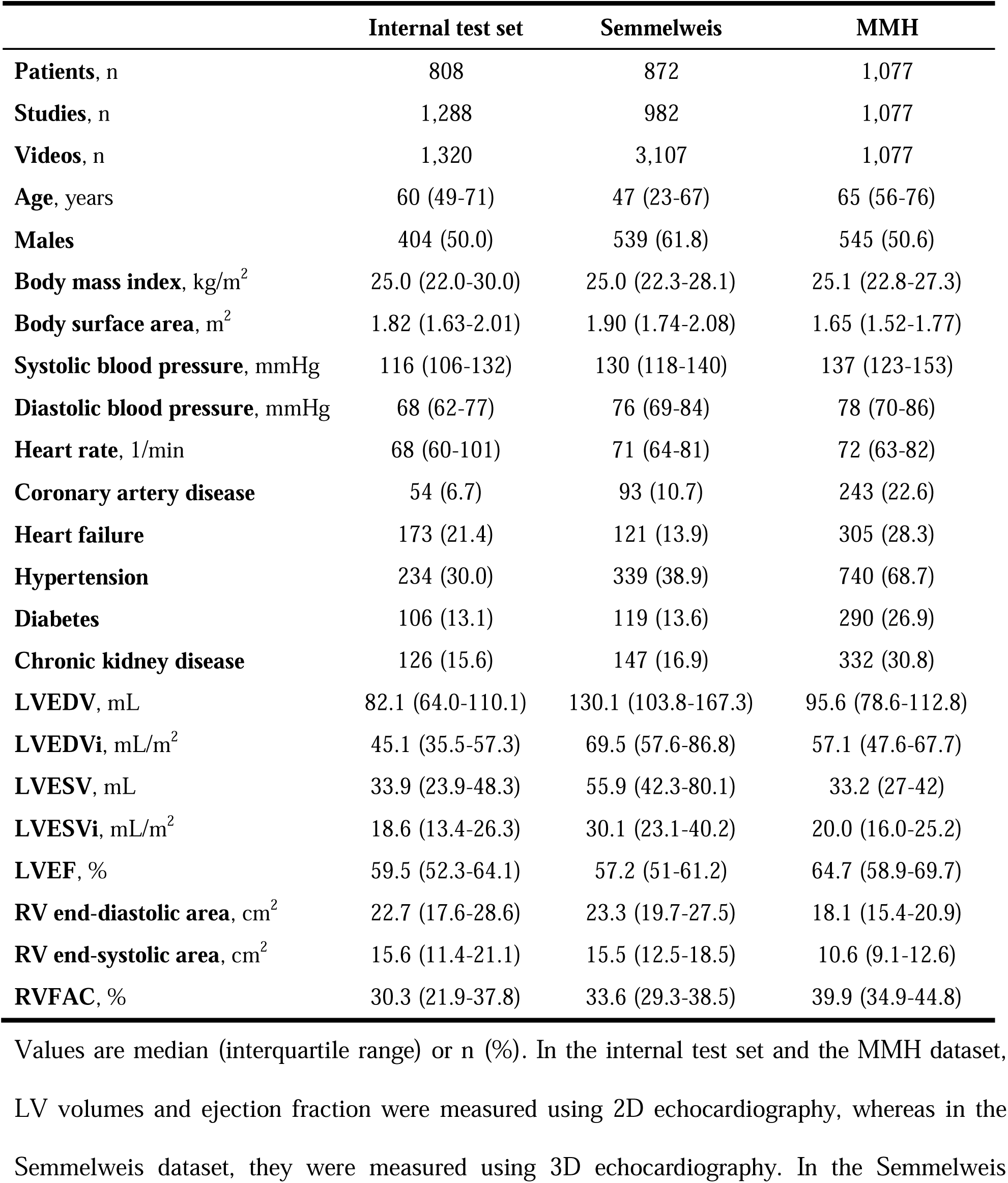
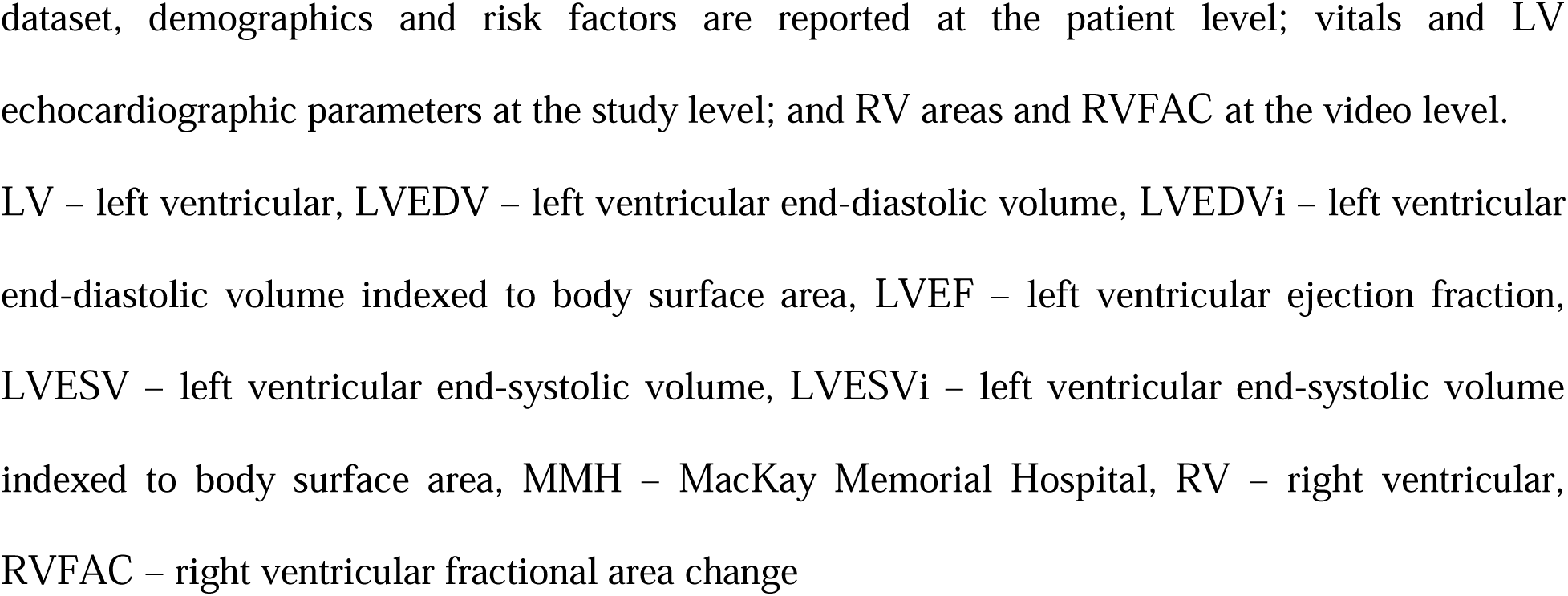
Clinical characteristics of the internal and external test sets.

|  | **Internal test set** | **Semmelweis** | **MMH** |
| --- | --- | --- | --- |
| **Patients, n** | 808 | 872 | 1,077 |
| **Studies, n** | 1,288 | 982 | 1,077 |
| **Videos, n** | 1,320 | 3,107 | 1,077 |
| **Age, years** | 60 (49-71) | 47 (23-67) | 65 (56-76) |
| **Males** | 404 (50.0) | 539 (61.8) | 545 (50.6) |
| **Body mass index, kg/m<sup>2</sup>** | 25.0 (22.0-30.0) | 25.0 (22.3-28.1) | 25.1 (22.8-27.3) |
| **Body surface area, m<sup>2</sup>** | 1.82 (1.63-2.01) | 1.90 (1.74-2.08) | 1.65 (1.52-1.77) |
| **Systolic blood pressure, mmHg** | 116 (106-132) | 130 (118-140) | 137 (123-153) |
| **Diastolic blood pressure, mmHg** | 68 (62-77) | 76 (69-84) | 78 (70-86) |
| **Heart rate, 1/min** | 68 (60-101) | 71 (64-81) | 72 (63-82) |
| **Coronary artery disease** | 54 (6.7) | 93 (10.7) | 243 (22.6) |
| **Heart failure** | 173 (21.4) | 121 (13.9) | 305 (28.3) |
| **Hypertension** | 234 (30.0) | 339 (38.9) | 740 (68.7) |
| **Diabetes** | 106 (13.1) | 119 (13.6) | 290 (26.9) |
| **Chronic kidney disease** | 126 (15.6) | 147 (16.9) | 332 (30.8) |
| **LVEDV, mL** | 82.1 (64.0-110.1) | 130.1 (103.8-167.3) | 95.6 (78.6-112.8) |
| **LVEDVi, mL/m<sup>2</sup>** | 45.1 (35.5-57.3) | 69.5 (57.6-86.8) | 57.1 (47.6-67.7) |
| **LVESV, mL** | 33.9 (23.9-48.3) | 55.9 (42.3-80.1) | 33.2 (27-42) |
| **LVESVi, mL/m<sup>2</sup>** | 18.6 (13.4-26.3) | 30.1 (23.1-40.2) | 20.0 (16.0-25.2) |
| **LVEF, %** | 59.5 (52.3-64.1) | 57.2 (51-61.2) | 64.7 (58.9-69.7) |
| **RV end-diastolic area, cm<sup>2</sup>** | 22.7 (17.6-28.6) | 23.3 (19.7-27.5) | 18.1 (15.4-20.9) |
| **RV end-systolic area, cm<sup>2</sup>** | 15.6 (11.4-21.1) | 15.5 (12.5-18.5) | 10.6 (9.1-12.6) |
| **RVFAC, %** | 30.3 (21.9-37.8) | 33.6 (29.3-38.5) | 39.9 (34.9-44.8) |

Values are median (interquartile range) or n (%). In the internal test set and the MMH dataset, LV volumes and ejection fraction were measured using 2D echocardiography, whereas in the Semmelweis dataset, they were measured using 3D echocardiography. In the Semmelweis
dataset, demographics and risk factors are reported at the patient level; vitals and LV echocardiographic parameters at the study level; and RV areas and RVFAC at the video level. LV – left ventricular, LVEDV – left ventricular end-diastolic volume, LVEDVi – left ventricular end-diastolic volume indexed to body surface area, LVEF – left ventricular ejection fraction, LVESV – left ventricular end-systolic volume, LVESVi – left ventricular end-systolic volume indexed to body surface area, MMH – MacKay Memorial Hospital, RV – right ventricular, RVFAC – right ventricular fractional area change

#### MacKay Memorial Hospital dataset

The MacKay Memorial Hospital (MMH) dataset, also used for external validation, included a total of 1,077 A4C videos from 1,077 studies of 1,077 patients who underwent transthoracic echocardiography between 2009 and 2022 at MacKay Memorial Hospital (Taipei, Taiwan). Clinical characteristics of these patients are summarized in Table 1, whereas technical information regarding the videos is provided in Supplemental Table 3.

### Video annotation

For each video in all three datasets, the endocardial border of the RV was manually traced by experienced echocardiographers in end-diastolic and end-systolic frames to assess the corresponding RV areas, which were then used to calculate RVFAC. In the Stanford dataset, manual contouring was performed in only one end-diastolic and one end-systolic frame per video, whereas in the two external test sets, contouring was performed in up to three cardiac cycles, when feasible. If contouring was performed in multiple cardiac cycles, RVFAC values were calculated using end-diastolic and end-systolic area pairs from each cardiac cycle and averaged to derive a single RVFAC label for the video. Of note, EchoNet-RV was trained using both the human expert-drawn RV contours (i.e., segmentation masks) of the sparsely annotated frames and the video-level RVFAC values. In addition, an experienced echocardiographer reviewed all videos of both external test sets to assess image quality using a 5-point Likert scale (nondiagnostic, poor, moderate, good, excellent) and to determine the view type (standard or RV-focused A4C), thereby enabling the analysis of these factors’ impact on model performance.

### Video preprocessing

An automated preprocessing pipeline was used to remove identifying information and burnt-in annotations, and to convert videos exported as DICOM files into a standardized format suitable for DL analysis. First, each video was square-cropped and masked to eliminate text and any other information outside the scanning sector. The resulting square frames were subsequently down-sampled to standardized 112×112-pixel videos using cubic interpolation and saved as AVI files.

### Architecture and development of EchoNet-RV

EchoNet-RV comprises two key modules, similar to EchoNet-Dynamic (13). The first module is a convolutional neural network (CNN) that uses a DeepLabV3 (22) architecture with a ResNet-50 backbone to perform frame-level semantic segmentation of the RV. Even though the data used for training this module was only sparsely annotated (i.e., human annotations were available for only some of the end-diastolic and end-systolic frames), the chosen architecture was well-suited for this form of weakly supervised learning, and generalized well throughout the cardiac cycle, even to frames that did not contain annotations (Figure 1A). The model was trained for 50 epochs using pixel-wise cross-entropy loss and a stochastic gradient descent optimizer with a learning rate of 1e-5, a momentum of 0.9, and a batch size of 20. The weights from the epoch with the lowest validation loss were selected for final testing.

**Figure 1.**
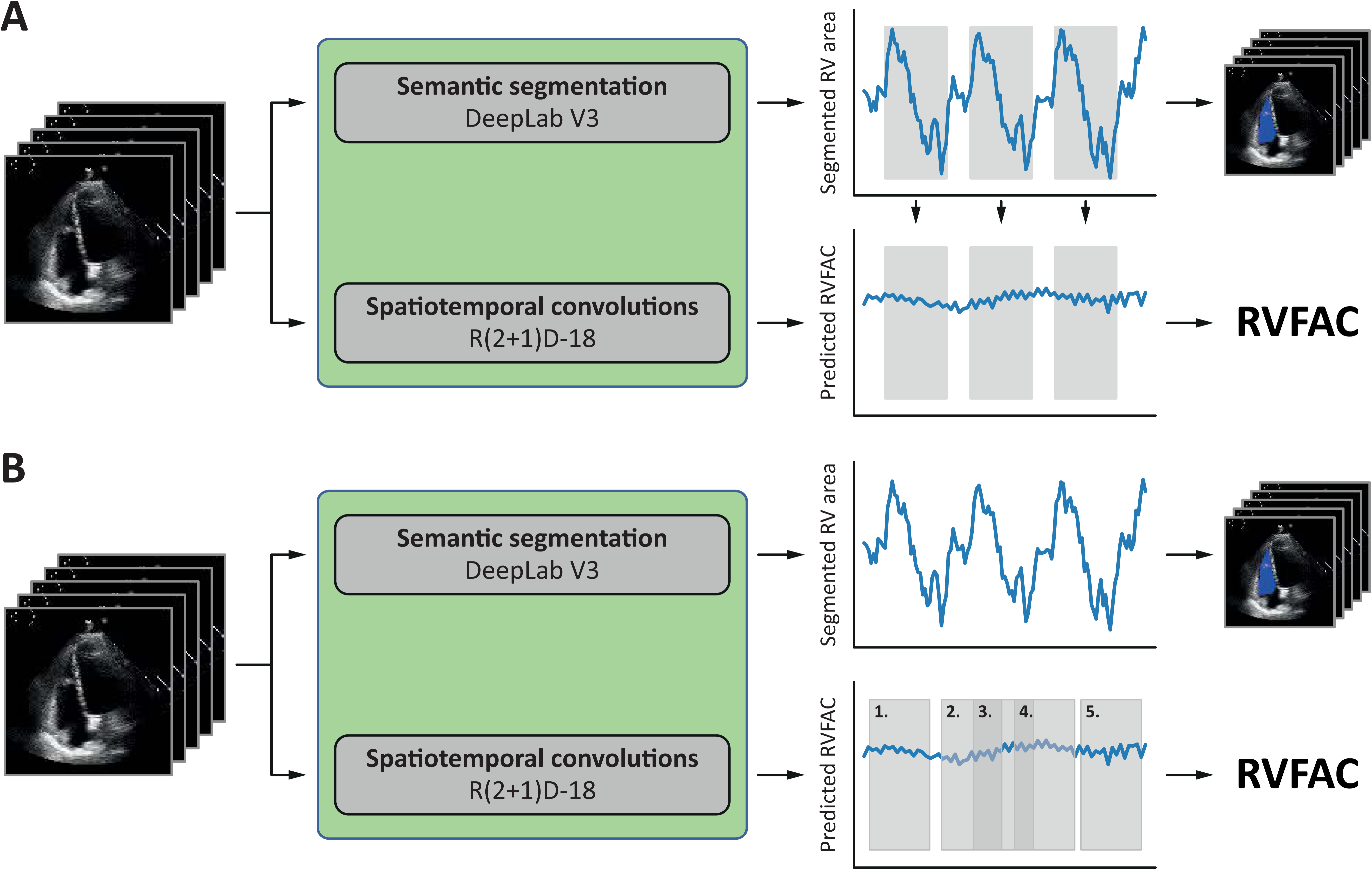
Methods and results of segmentation. (A) Weakly supervised training of the semantic segmentation module using human expert tracings of RV areas on ED and ES frames and both annotated and non-annotated frames of the input video. (B) Segmented RV areas for all frames of a video from a patient in sinus rhythm and another from a patient in atrial fibrillation. (C) Performance of EchoNet-RV in segmenting the RV in the three test sets. ED – end-diastolic, ES – end-systolic, MMH – MacKay Memorial Hospital, RV – right ventricular

The second module is a spatiotemporal CNN with an R(2+1)D-18 (23) architecture that directly estimates RVFAC from each video without relying on segmenting the RV. During the training of this module, 16 frames were sampled by taking every other frame from the video as input. To simulate slight translations and changes in transducer position, training videos were padded by 12 pixels on each side, followed by a random crop of the original frame size. The model was trained for 45 epochs using a stochastic gradient descent optimizer with a learning rate of 1e-4, a momentum of 0.9, and a batch size of 20.

Given that substantial beat-to-beat variation in the end-diastolic and end-systolic RV areas (and thus in RVFAC) can occur in several cardiac conditions, such as atrial fibrillation and premature atrial or ventricular contractions, test-time augmentation was also applied to obtain robust final predictions. Experiments were conducted using two different approaches (Figure 2). The first approach followed the method used in EchoNet-Dynamic (13). Briefly, each ventricular contraction (i.e., the systolic phase of the cardiac cycle) was identified by selecting the frames between the largest and smallest RV areas predicted by the segmentation model. For each identified contraction (i.e., beat), 32-frame video clips centred around the ventricular contraction were obtained and processed by the spatiotemporal CNN module to produce beat-to-beat estimates of RVFAC, which were then averaged to yield the final video-level output of EchoNet-RV. In contrast, the second approach did not rely on the segmentation model’s output. Instead, it randomly sampled 5 (possibly overlapping) 32-frame video clips from each video, and individual predictions generated by the spatiotemporal CNN module from these 5 clips were averaged to produce the final video-level model output. The approach achieving superior performance in the held-out test set was adopted as the ultimate test-time augmentation technique in the final EchoNet-RV model, and the results are reported accordingly.

**Figure 2.**
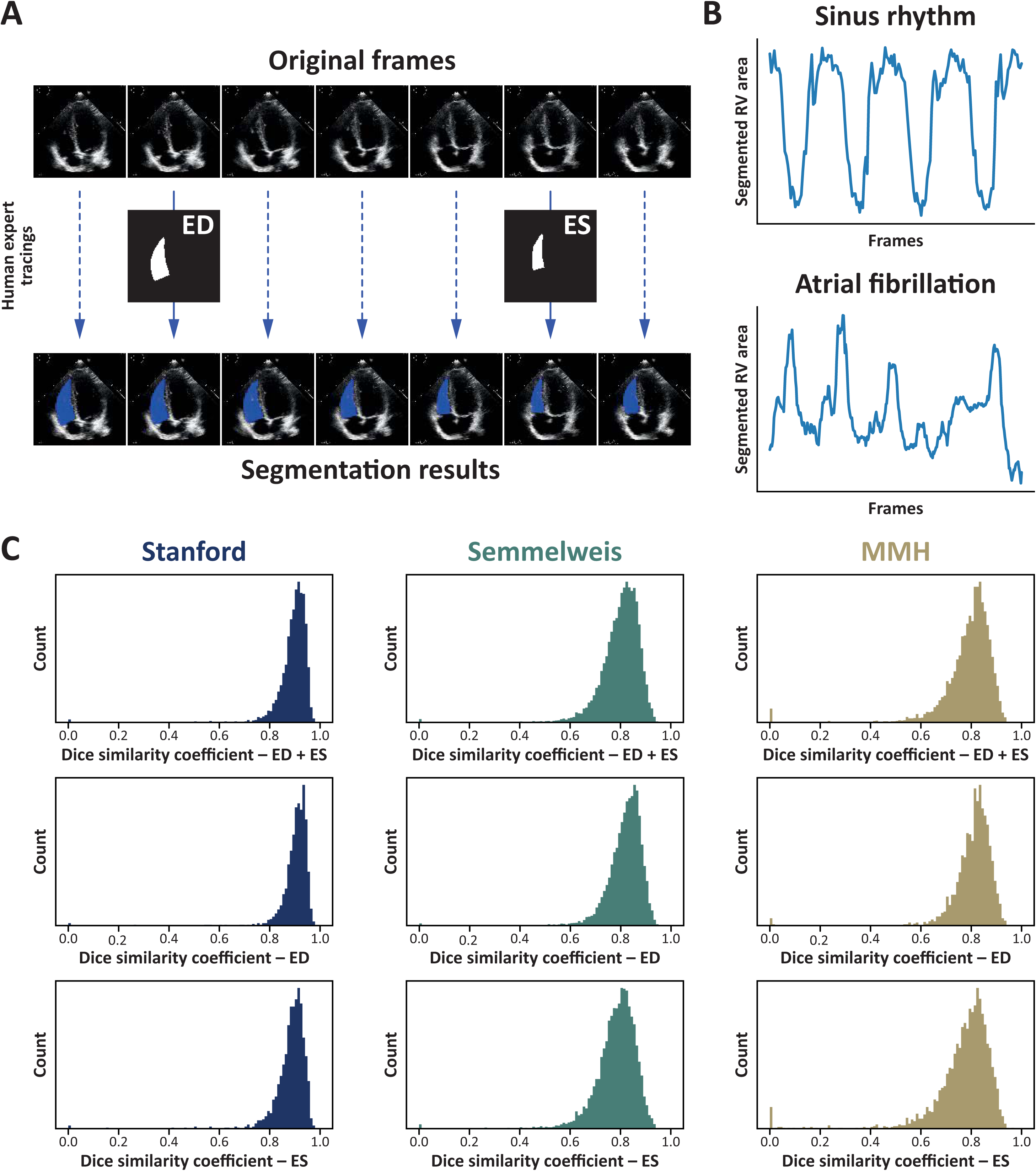
Schematic illustration of the two test-time augmentation strategies tested in the study. (A) In the first approach, each ventricular contraction was identified by selecting the frames between the largest and smallest RV areas predicted by the semantic segmentation module. For each identified contraction (i.e., beat), 32-frame video clips centred around the ventricular contraction were obtained and processed by the spatiotemporal CNN module to produce **beat-to-beat** estimates of RVFAC, which were then averaged to yield the final video-level output of EchoNet-RV. (B) The second approach did not rely on the segmentation model’s outputs. Instead, 5 (possibly overlapping) 32-frame video clips were **randomly sampled** from each video, and individual predictions generated by the spatiotemporal CNN module from these 5 clips were averaged to produce the final video-level model output. CNN – convolutional neural network, RVFAC – right ventricular fractional area change; other abbreviations as in Figure 1.

Preprocessing and model development were performed in Python (version 3.15) using the PyTorch DL library (version 1.8.0). Models were trained on a server equipped with four NVIDIA GeForce RTX 2080 Ti graphics processing units. To facilitate further research and potential deployment, the public repository containing the source code and model weights of EchoNet-RV is available at https://github.com/echonet/RV.

### Comparing prediction error with beat-to-beat and inter-observer variability

To compare EchoNet-RV’s prediction error with beat-to-beat variability, videos of the external test sets with human measurements of RVFAC in 2 or more cardiac cycles (n=3,943) were used. To compare the model’s prediction error with inter-observer variability, 250 videos from each of the Semmelweis and MMH datasets were randomly sampled, and RVFAC was remeasured by a second reader blinded to the first reader’s measurements. These repeated measurements were used for quantifying prediction error, beat-to-beat variability, and inter-observer variability. Metrics and statistical tests used in the analyses are described in the subsection on performance metrics.

### Benchmarking against multi-task models

EchoNet-RV’s performance was also compared with two recently published multi-task DL models for comprehensive echocardiogram interpretation – EchoPrime (21) and PanEcho (20) – in the external test sets. While EchoPrime estimates RVFAC as a continuous variable, PanEcho can only classify RV systolic function as normal or reduced. Therefore, the performance of EchoNet-RV and EchoPrime could be compared both in estimating RVFAC and in detecting RV dysfunction, whereas benchmarking against PanEcho was possible only for the latter task.

### Analyzing associations with outcomes

Associations between the predicted RVFAC values and outcomes were analyzed in the Semmelweis dataset. In the present study, the outcomes of interest were the composite endpoint of heart failure hospitalization or all-cause death, as well as all-cause death alone.

### Performance metrics and statistical analysis

EchoNet-RV’s performance in the semantic segmentation task was evaluated using the Dice similarity coefficient, whereas its performance in the RVFAC estimation task was assessed using the mean absolute error (MAE) and the intra-class correlation coefficient (ICC). Absolute errors between subgroups were compared using the unpaired Student’s t-test or the Mann-Whitney U test, as appropriate. The trend between image quality categories and absolute errors was assessed using the Jonckheere-Terpstra test. Bland-Altman analysis was performed to calculate the bias and limits of agreement (LOA). Additionally, an extended version of the Bland-Altman analysis, incorporating inverse weighting, defining a clinically relevant range of interest, and quantifying the trend between means and differences, was also performed, as proposed by Pasdeloup et al. (24). In the present study, the range of interest for RVFAC was 15–45%, corresponding to the clinically relevant spectrum observed across the datasets. The model’s performance in detecting RV dysfunction (defined as RVFAC <35%) was quantified using the area under the receiver operating characteristic curve (AUC), and accuracy, specificity, sensitivity, negative predictive value, and positive predictive value were also computed after dichotomizing the predicted and human-measured RVFAC values using the guideline-recommended cutoff value of 35% (9). To evaluate EchoNet-RV’s prediction error in the context of beat-to-beat and inter-observer variability, mean absolute difference (MAD), standard error of measurement (SEM), minimal detectable change (MDC), and coefficient of variation (CV) were computed in subsets of videos. MADs were compared using paired Student’s t-tests or Wilcoxon signed-rank tests, whereas the significance of the difference in SEM, MDC, and CV was determined using bootstrap p-values. When benchmarking the performance of EchoNet-RV against other models, absolute errors were compared using paired Student’s t-tests or Wilcoxon signed-rank tests, whereas AUCs were compared using the DeLong test. For each of the aforementioned performance metrics, 95% confidence intervals (CIs) were calculated from 10,000 stratified bootstrap resamples. Results are reported at the video level unless stated otherwise.

Continuous variables are expressed as median (interquartile range), while categorical variables are reported as frequencies and percentages. The event-free survival of subgroups was visualized on Kaplan-Meier curves, and Log-rank tests were performed for comparison. In patients with follow-up longer than 5 years and no events during that period, right censoring was applied at 5 years. Cox proportional hazards models were used to compute hazard ratios with 95% CIs. All survival analyses were performed at the patient level. A p-value of <0.05 was considered statistically significant.

### Ethical approval

The study protocol conforms with the principles outlined in the Declaration of Helsinki, and it was approved by the Institutional Review Boards of Stanford University and Cedars-Sinai Medical Center, the Regional and Institutional Committee of Science and Research Ethics of Semmelweis University (approval number: 190/2020), and the Institutional Review Board of MacKay Memorial Hospital (study identifier: 25MMHIS019e). Obtaining informed consent was waived due to the retrospective nature of the analysis. Methods and results are reported in compliance with the updated Proposed Requirements for Cardiovascular Imaging-Related Multimodal-AI Evaluation (PRIME 2.0) checklist (Supplemental Table 4) (25).

## RESULTS

### Performance in RV segmentation

EchoNet-RV segmented the RV in all human-annotated end-diastolic and end-systolic frames with Dice coefficients of 0.893 (95% CI: 0.891–0.895), 0.797 (95% CI: 0.795–0.798), and 0.788 (95% CI: 0.785–0.790) in the held-out internal test set and Semmelweis and MMH datasets, respectively (Figure 1B–C). When considering only end-diastolic frames, it achieved Dice coefficients of 0.903 (95% CI: 0.900–0.905), 0.815 (95% CI: 0.813–0.816), and 0.807 (95% CI: 0.805–0.810) in these three datasets, whereas in end-systolic frames only, the Dice coefficients were 0.883 (95% CI: 0.880–0.886), 0.779 (95% CI: 0.777–0.781), and 0.768 (95% CI: 0.764–0.772), respectively (Figure 1B–C).

### Performance in RVFAC prediction

Among the two evaluated test-time augmentation strategies, random sampling of 5 clips yielded a slightly lower MAE than beat-to-beat sampling (5.795 [95% CI: 5.520–6.070] vs. 5.833 [95% CI: 5.555–6.113] percentage points) in the held-out test set; therefore, this approach was adopted in the final EchoNet-RV model.

EchoNet-RV predicted RVFAC with MAEs of 5.795 (95% CI: 5.560–6.031), 5.830 (95% CI: 5.692–5.970), and 6.362 (95% CI: 6.064–6.660) percentage points and ICCs of 0.648 (95% CI: 0.616–0.677), 0.481 (95% CI: 0.452–0.509), and 0.301 (95% CI: 0.243–0.356) in the held-out test set and the Semmelweis and MMH datasets, respectively (Figure 3A). Conventional Bland-Altman analysis showed biases of 0.233, 1.275, and -2.635 percentage points, along with LOA widths of 30.260, 28.837, and 29.987 percentage points, in the three test sets (Figure 3B). Extended Bland-Altman analysis, which is less affected by the distribution differences between datasets (24,26), showed biases of 0.433, 1.875, and -0.228 percentage points, LOA widths of 29.765, 31.221, and 29.651 percentage points, and slopes of the mean-difference relationship of - 0.318, -0.210, and -0.354, respectively (Figure 3C). Given that multiple A4C videos were extracted from the same echocardiographic study in the Semmelweis dataset, study-level MAE and ICC were also calculated, yielding 4.744 (95% CI: 4.541–4.943) percentage points and 0.573 (95% CI: 0.526–0.617), respectively.

**Figure 3.**
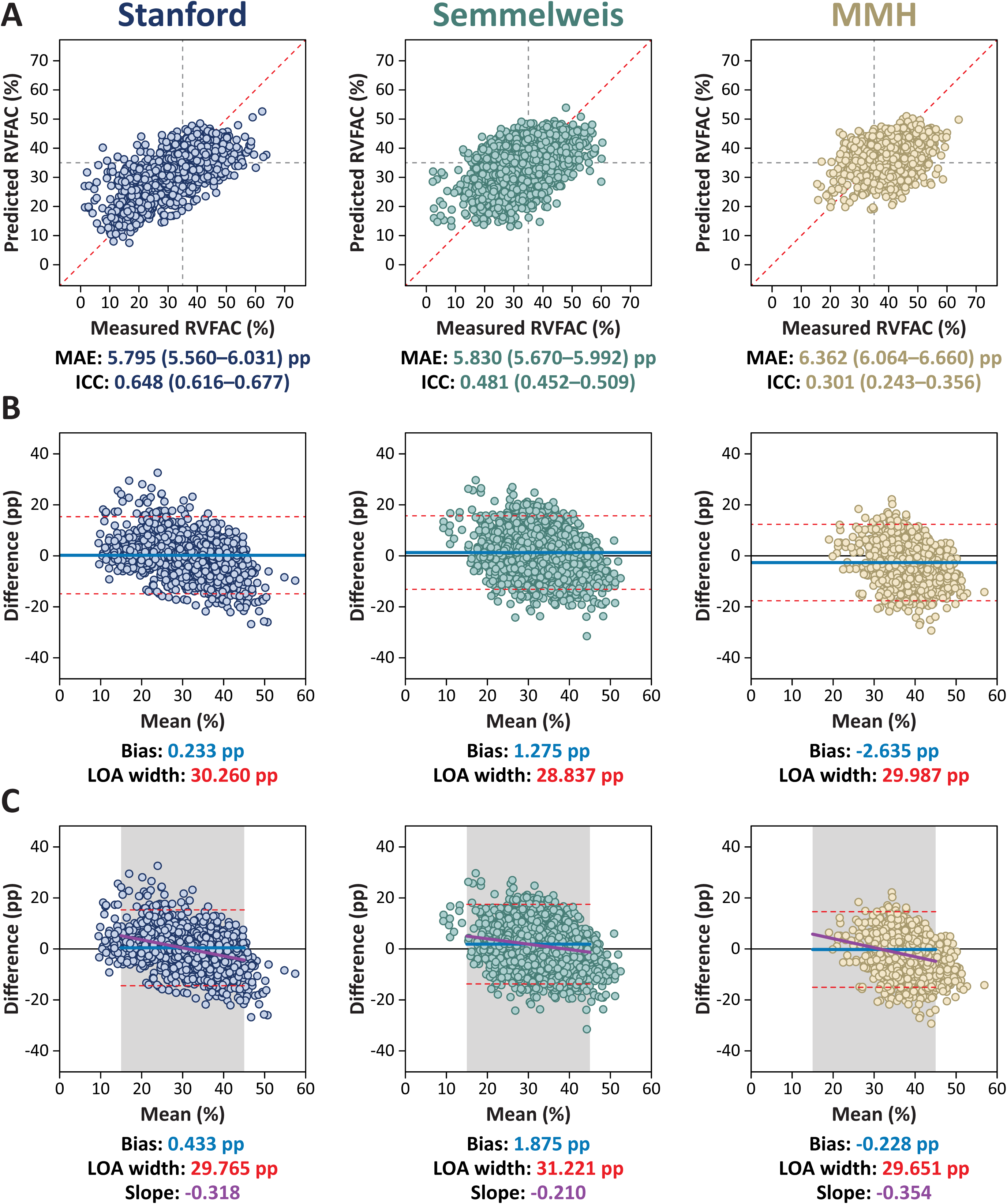
Performance of EchoNet-RV in predicting RVFAC. (A) Scatter plots showing the predicted vs. human-measured RVFAC values. (B) Conventional Bland-Altman plots showing the agreement between the predicted and human-measured RVFAC values. (C) Results of the extended Bland-Altman analysis that incorporates inverse weighting, defining a clinically relevant range of interest (RVFAC of 15–45%), and quantifying the trend between means and differences. ICC – intraclass correlation coefficient, LOA – limit of agreement, MAE – mean absolute error, pp – percentage points; other abbreviations as in Figures 1 and 2.

Although EchoNet-RV’s prediction error was higher than the beat-to-beat variability of a reader’s measurements within the same video (all p<0.001, Table 2), it was lower than the inter-observer variability (all p<0.001, Table 3).

**Table 2.** Comparison of EchoNet-RV’s prediction error with the beat-to-beat variability of human measurements in the external test sets.

|  | **EchoNet-RV** | **Beat-to-beat** | **P-value** |
| --- | --- | --- | --- |
| **Mean absolute difference, pp** | 5.874<br>(5.729-6.021) | 3.511<br>(3.409-3.612) | <0.001 |
| **Standard error of measurement, pp** | 3.746<br>(3.657-3.835) | 2.130<br>(2.066-2.196) | <0.001 |
| **Minimal detectable change, pp** | 10.384<br>(10.137-10.631) | 5.904<br>(5.727-6.088) | <0.001 |
| **Coefficient of variation, %** | 14.956<br>(14.598-15.326) | 10.132<br>(9.826-10.448) | <0.001 |

This analysis only included videos (n=3,943) with RVFAC measurements in 2 or more cardiac cycles. For each metric, 95% confidence intervals were calculated from 10,000 stratified bootstrap resamples. Mean absolute differences were compared using paired Student's t-test or Wilcoxon signed-rank test, whereas the significance of the difference in standard error of measurement, minimal detectable change, and coefficient of variation was determined using bootstrap p-values.
Abbreviations as in Table 1.

**Table 3.** Comparison of EchoNet-RV’s prediction error with the inter-reader variability of human measurements in the external test sets.

|  | **EchoNet-RV** | **Inter-reader** | **P-value** |
| --- | --- | --- | --- |
| **Mean absolute difference, pp** | 6.126<br>(5.735–6.563) | 9.699<br>(9.031–10.458) | <0.001 |
| **Standard error of measurement, pp** | 3.063<br>(2.868–3.282) | 4.849<br>(4.515–5.229) | <0.001 |
| **Minimal detectable change, pp** | 8.490<br>(7.948–9.096) | 13.442<br>(12.516–14.495) | <0.001 |
| **Coefficient of variation, %** | 12.568<br>(11.713–13.573) | 17.651<br>(16.475–19.015) | <0.001 |

This analysis included 250-250 randomly selected videos from the MMH and Semmelweis datasets. In each of these 500 videos, RVFAC was remeasured by a second reader blinded to the first reader’s measurements. For each metric, 95% confidence intervals were calculated from 10,000 stratified bootstrap resamples. Mean absolute differences were compared using paired Student’s t-test or Wilcoxon signed-rank test, whereas the significance of the difference in standard error of measurement, minimal detectable change, and coefficient of variation was determined using bootstrap p-values.
Abbreviations as in Table 1.

In the external test sets, subgroup analyses were also performed to analyze the differences in performance between videos acquired using different ultrasound machines and the impact of view type and image quality on prediction performance (Figure 4, Supplemental Table 5). These analyses revealed that the EchoNet-RV performed slightly better on videos acquired using GE ultrasound machines than those acquired using Philips (Semmelweis dataset: p<0.001; MMH dataset: p=0.031). Errors were similar in standard and RV-focused A4C videos in both external test sets (Semmelweis dataset: p=0.406; MMH dataset: p=0.072), whereas they showed a decreasing trend with better image quality in the Semmelweis dataset (p<0.001), but not in the MMH dataset (p=0.373).

**Figure 4.**
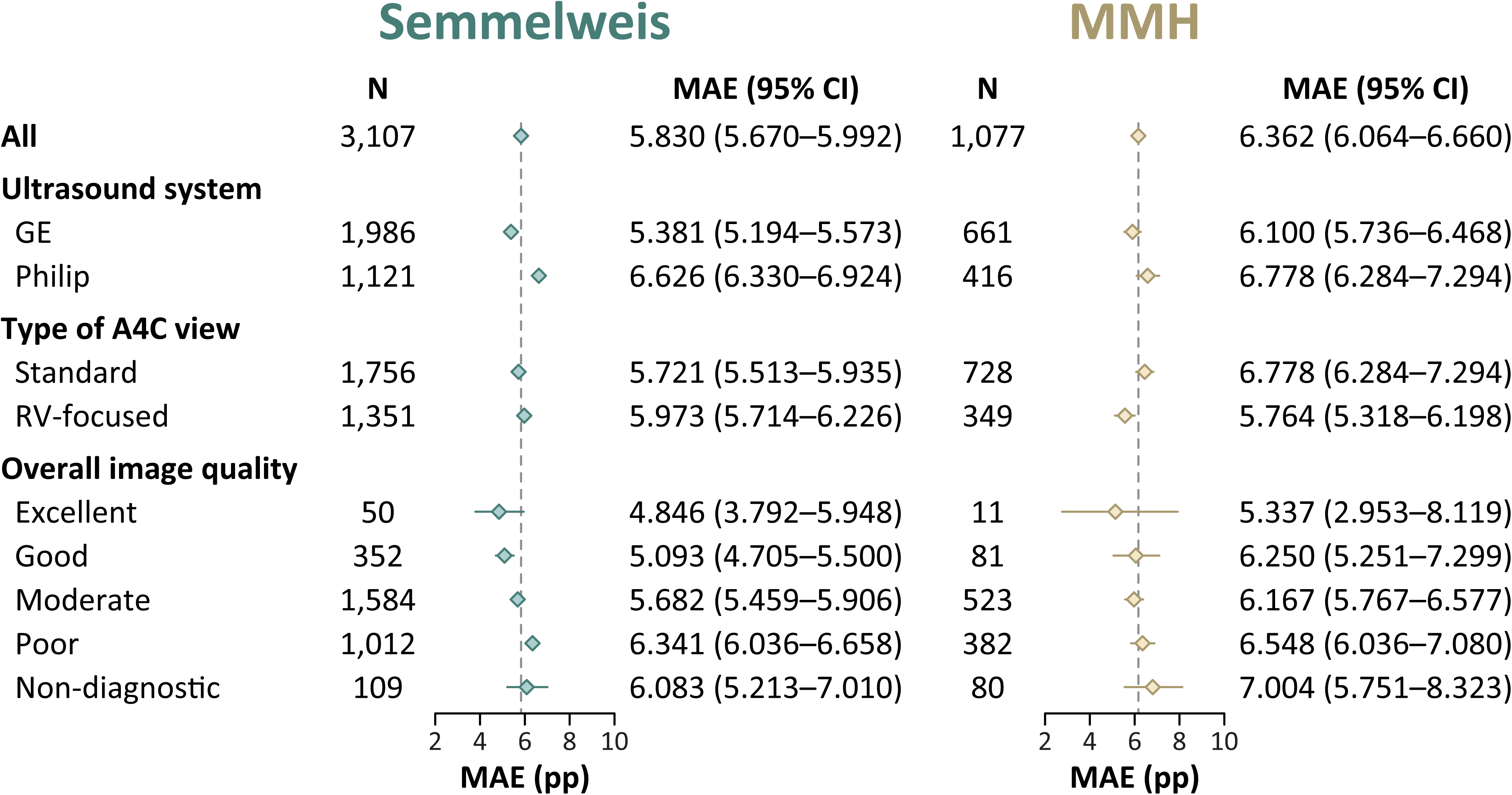
Performance of EchoNet-RV in predicting RVFAC in different subgroups of the external test sets. A4C – apical 4-chamber view; other abbreviations as in Figures 1 and 3.

### Performance in identifying RV dysfunction

RV dysfunction (defined as RVFAC <35%) was present in 878 (66.5%), 1,732 (55.7%), and 275 (25.5%) videos in the held-out test set and the Semmelweis and MMH datasets, respectively. EchoNet-RV identified RV dysfunction with AUCs of 0.859 (95% CI: 0.843–0.876), 0.725 (95% CI: 0.710–0.740), and 0.684 (95% CI: 0.653–0.713) in these three test sets, respectively (Figure 5). In the Semmelweis dataset, study-level AUC was 0.770 (95% CI: 0.746–0.794). Other performance metrics for this task are reported in Supplemental Table 6.

**Figure 5.**
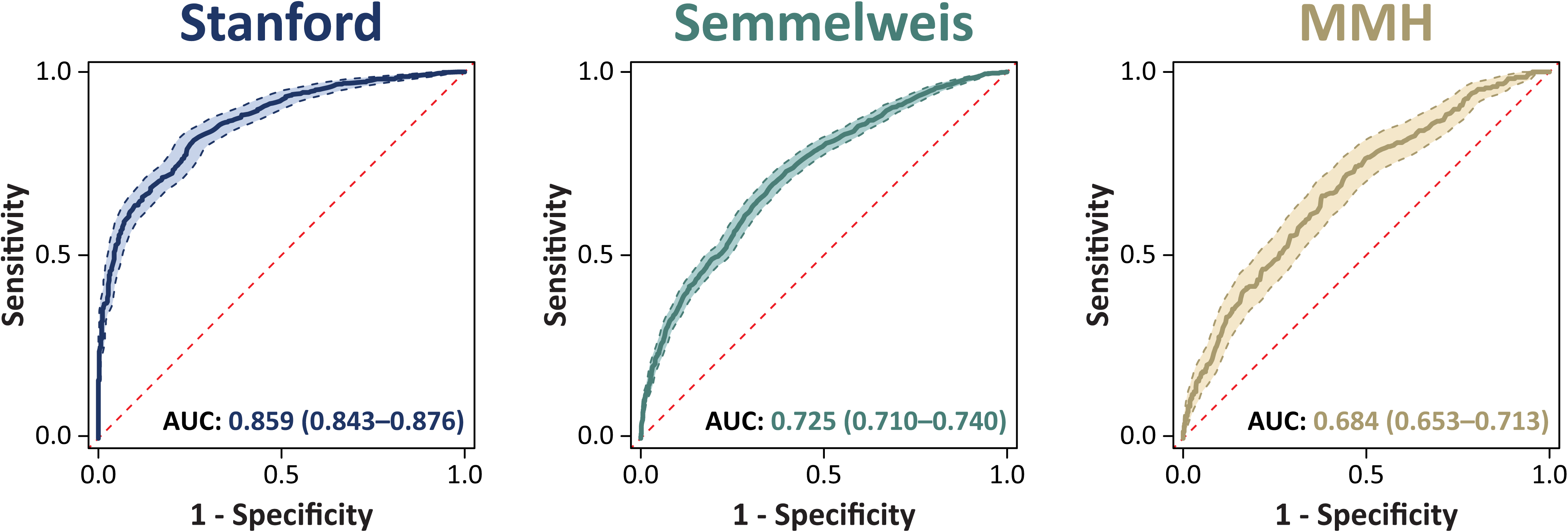
Receiver operating characteristic curves illustrating the performance of EchoNet-RV in identifying RV dysfunction. AUC – area under the receiver operating characteristic curve; other abbreviations as in Figure 1.

### Benchmarking against multi-task models

EchoNet-RV predicted RVFAC with lower absolute errors than EchoPrime in both the Semmelweis (5.830 [95% CI: 5.670–5.992] vs. 6.171 [95% CI: 6.002–6.339], p=0.001) and MMH datasets (6.362 [95% CI: 6.064–6.660] vs. 6.728 [95% CI: 6.406–7.048], p=0.009) (Table 4). Moreover, EchoNet-RV achieved higher AUCs for identifying RV dysfunction than both EchoPrime and PanEcho in the Semmelweis dataset (0.727 [95% CI: 0.709–0.744] vs. 0.683 [95% CI: 0.664–0.701] and vs. 0.673 [95% CI: 0.654–0.692], both p<0.001), whereas comparable AUCs were observed in the MMH dataset (0.684 [95% CI: 0.648–0.719] vs. 0.652 [95% CI: 0.614–0.690] and vs. 0.657 [95% CI: 0.619–0.695], p=0.220 and p=0.150, respectively) (Table 4).

**Table 4.** Performance of EchoNet-RV, EchoPrime, and PanEcho in the external test sets.

|  | **EchoNet-RV** | **EchoPrime** | **PanEcho** |
| --- | --- | --- | --- |
| **Semmelweis (video-level)** |  |  |  |
| MAE, pp | 5.830<br>(5.670–5.992) | 6.171<br>(6.002–6.339) | - |
| ICC | 0.481<br>(0.452–0.509) | 0.423<br>(0.394–0.450) | - |
| AUC | 0.727<br>(0.709–0.744) | 0.683<br>(0.664–0.701) | 0.673<br>(0.654–0.692) |
| **Semmelweis (study-level)** |  |  |  |
| MAE, pp | 4.744<br>(4.541–4.943) | 5.475<br>(5.214–5.743) | - |
| ICC | 0.573<br>(0.526–0.617) | 0.488<br>(0.437–0.534) | - |
| AUC | 0.770<br>(0.746–0.794) | 0.714<br>(0.681–0.745) | 0.687<br>(0.653–0.720) |
| **MMH** |  |  |  |
| MAE, pp | 6.362<br>(6.064–6.660) | 6.728<br>(6.406–7.048) | - |
| ICC | 0.301<br>(0.243–0.356) | 0.192<br>(0.127–0.253) | - |
| AUC | 0.684<br>(0.648–0.719) | 0.652<br>(0.614–0.690) | 0.657<br>(0.619–0.695) |

Performance metrics are reported with their corresponding 95% confidence intervals computed using 10,000 bootstrapped samples. AUCs are reported for identifying videos/studies with an actual RVFAC of less than 35%.

### Associations with outcomes

In the Semmelweis dataset, 125 (14.3%) patients reached the composite endpoint of heart failure hospitalization or all-cause death, whereas 110 (12.6%) died during the follow-up duration of 5.0 (4.2–5.0) years. Univariable Cox regression analyses showed that the higher values of EchoNet-RV-predicted RVFAC were associated with a lower risk of the composite endpoint (undajusted HR: 0.877 [95% CI: 0.855–0.900], p<0.001) and all-cause death (undajusted HR: 0.877 [95% CI: 0.854–0.901], p<0.001) (Tables 5 and 6). Moreover, predicted RVFAC was also an independent predictor of these adverse outcomes in multivariable Cox regression models (composite endpoint – adjusted HR: 0.943 [95% CI: 0.912–0.975], p<0.001; all-cause death – adjusted HR: 0.932 [95% CI: 0.899–0.965], p<0.001) that included age, sex, coronary artery disease, heart failure, hypertension, diabetes, chronic kidney disease, and left ventricular ejection fraction as covariates (Tables 5 and 6). The prognostic value of the model’s predictions was also confirmed by Kaplan-Meier curves of subgroups created based on the guideline-recommended RVFAC cutoff value of 35% (Figure 6).

**Figure 6.**
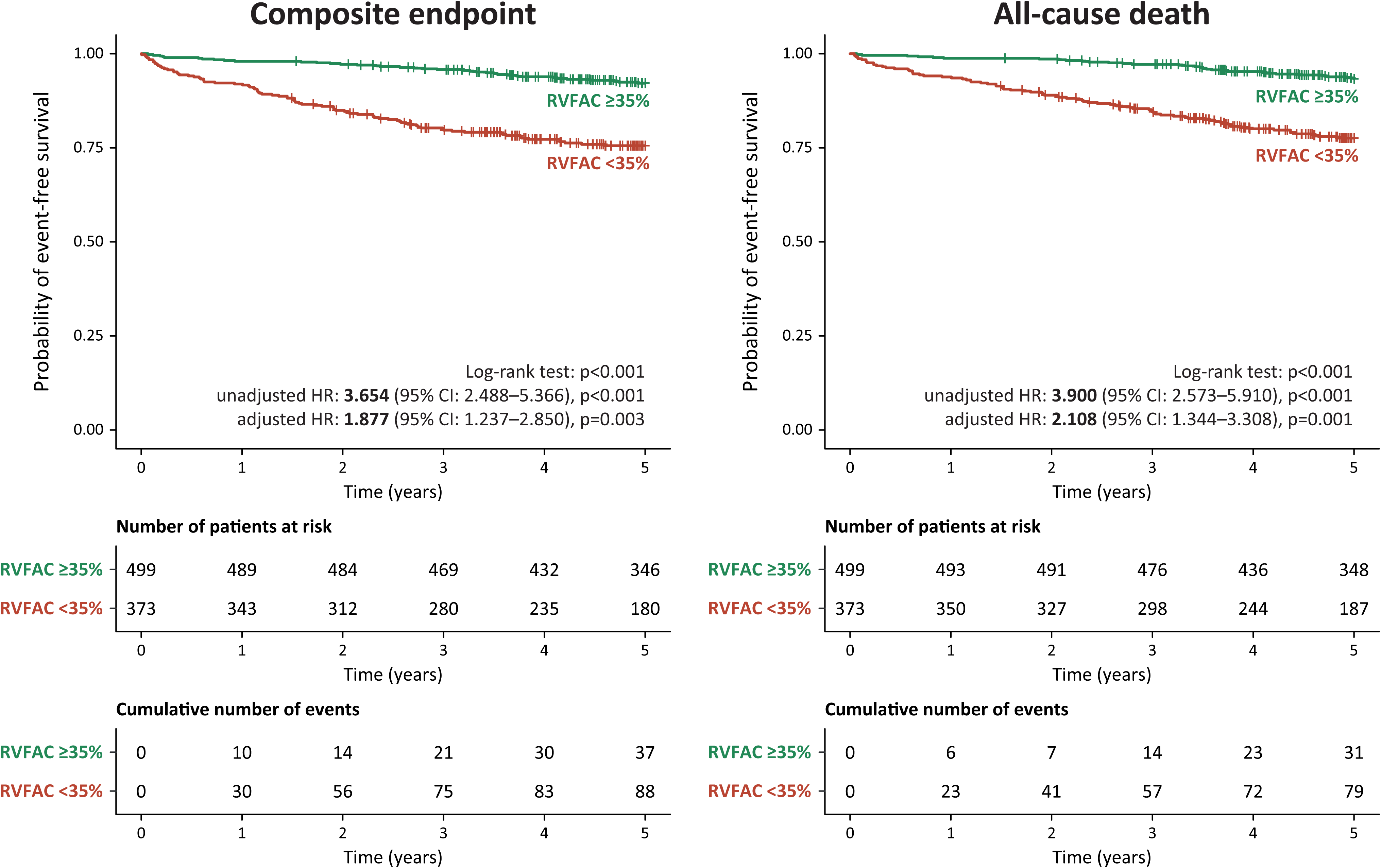
Kaplan-Meier curves showing the event-free survival of patient subgroups in the Semmelweis dataset. Abbreviations as in Figure 2.

**Table 5.** Associations between the EchoNet-RV-predicted RVFAC values and the composite of heart failure hospitalization or all-cause death.

|  | **Univariable Cox regression** |  | **Multivariable Cox regression** |  |
| --- | --- | --- | --- | --- |
|  | HR (95% CI) | P-value | HR (95% CI) | P-value |
| Age | 1.072 (1.059–1.085) | <0.001 | 1.060 (1.043–1.077) | <0.001 |
| Male sex | 1.528 (1.041–2.244) | 0.030 | 1.059 (0.703–1.595) | 0.783 |
| Coronary artery disease | 4.204 (3.197–5.528) | <0.001 | 1.257 (0.896–1.762) | 0.185 |
| Heart failure | 6.980 (4.876–9.992) | <0.001 | 1.301 (0.725–2.336) | 0.377 |
| Hypertension | 6.943(4.529–10.644) | <0.001 | 1.155 (0.701–1.904) | 0.571 |
| Diabetes | 4.871 (3.385–7.009) | <0.001 | 1.598 (1.086–2.350) | 0.017 |
| Chronic kidney disease | 1.669 (1.112–2.505) | 0.013 | 0.741 (0.483–1.136) | 0.169 |
| LVEF | 0.936 (0.926–0.946) | <0.001 | 0.990 (0.969–1.011) | 0.342 |
| EchoNet-RV-predicted RVFAC | 0.877 (0.855–0.900) | <0.001 | 0.943 (0.912–0.975) | <0.001 |

CI – confidence interval, HR – hazard ratio; other abbreviations as in Table 1.

**Table 6.** Associations between the EchoNet-RV-predicted RVFAC values and all-cause death.

|  | Univariable Cox regression |  | Multivariable Cox regression |  |
| --- | --- | --- | --- | --- |
|  | HR (95% CI) | P-value | HR (95% CI) | P-value |
| Age | 1.074 (1.059–1.088) | <0.001 | 1.061 (1.043–1.079) | <0.001 |
| Male sex | 1.567 (1.039–2.365) | 0.032 | 1.076 (0.695–1.666) | 0.742 |
| Coronary artery disease | 4.256 (3.208–5.647) | <0.001 | 1.364 (0.964–1.932) | 0.080 |
| Heart failure | 6.744 (4.609–9.868) | <0.001 | 1.508 (0.790–2.879) | 0.213 |
| Hypertension | 7.319 (4.616–11.606) | <0.001 | 1.237 (0.725–2.111) | 0.434 |
| Diabetes | 4.931 (3.348–7.263) | <0.001 | 1.546 (1.029–2.325) | 0.036 |
| Chronic kidney disease | 1.923 (1.264–2.926) | 0.002 | 0.912 (0.586–1.419) | 0.683 |
| LVEF | 0.940 (0.929–0.951) | <0.001 | 1.003 (0.980–1.026) | 0.801 |
| EchoNet-RV-predicted RVFAC | 0.877 (0.854–0.901) | <0.001 | 0.932 (0.899–0.965) | <0.001 |

CI – confidence interval, HR – hazard ratio; other abbreviations as in Table 1.

## DISCUSSION

The main findings of our study can be summarized as follows: (i) we developed and internally validated EchoNet-RV, a DL model capable of accurately asessing RVFAC and identifying RV dysfunction from A4C echocardiographic videos, (ii) we externally validated EchoNet-RV’s performance in two large datasets, confirming the model’s generalizability across diverse patient populations, geographic regions, ultrasound machine vendors, and image qualities, (iii) we demonstrated that EchoNet-RV’s prediction errors are lower than the inter-observer variability observed among human experts, (iv) we showed that, as a single-task model, EchoNet-RV can outperform multi-task models in estimating RVFAC and indetifying RV dysfunction, and (v) we found that the predicted RVFAC values are significantly and independently associated with adverse clinical outcomes. Additionally, we released the code and model weights of EchoNet-RV to promote further research and facilitate deployment in other research or clinical environments.

AI has the potential to substantially enhance cardiac imaging workflows, particularly the challenging echocardiographic evaluation of RV function (7,27). Variability in acquisition protocols and technical settings, image analysis, and interpretation of RV parameters remains a major obstacle to the consistent clinical assessment of RV function across institutions and geographic regions (3). Standardized measurements are essential not only for accurate diagnosis and disease monitoring but also for guiding therapeutic decisions and ensuring reliable longitudinal follow-up. In this context, AI could be a valuable complement even to conventional core laboratory workflows by providing reproducible, objective, and scalable solutions for automating measurements and echocardiogram interpretation (28). While current commercially available AI tools for echocardiography support automated analysis of a limited set of RV parameters, more advanced metrics and broader implementation and validation across diverse clinical environments remain necessary to fully realize the benefits of AI-driven RV function assessment (17,29). To date, no publicly available tool has been proposed that enables vendor-independent and consistent evaluation of RVFAC.

Despite their ease of use and reproducibility, the most recent American Society of Echocardiography guidelines emphasize the importance of not relying solely on measures of RV longitudinal shortening, such as TAPSE and S’ (9). In specific clinical scenarios, such as following open-heart surgery or in patients with left ventricular systolic dysfunction, the mechanical pattern of RV contractions can shift toward a more radial orientation, and parameters quantifying solely the longitudinal function may underestimate global RV function (30,31). Conversely, in conditions characterized by RV pressure overload, longitudinal function often remains preserved in the initial phase of the disease, while radial contraction is the first to deteriorate (32). Although RVFAC is the most comprehensive two-dimensional parameter for assessing RV function, as it captures both longitudinal and radial shortening, as well as the contribution of the interventricular septum, it remains only the third most commonly adopted quantitative method for RV assessment, primarily due to its relatively limited reproducibility and the time-consuming nature of its measurement in routine clinical practice (3,4). Consistent with previous studies, we observed substantial inter-observer variability in RVFAC measurements, even among experienced readers. Notably, the prediction errors of EchoNet-RV compared to human measurements were lower than the inter-observer variability among human readers. While the model demonstrated acceptable discriminative power in differentiating normal RV function from RV dysfunction using a 35% RVFAC threshold, it is important to highlight that EchoNet-RV performs regression and estimates RVFAC on a continuous scale. This approach allows for more granular assessment of RV function and may facilitate classification across the spectrum of dysfunction severity, aligning with recent guideline recommendations (9). Furthermore, the predicted RVFAC values were independently associated with both all-cause death and a composite endpoint of heart failure hospitalization or all-cause death, even after adjustment for relevant clinical covariates, including age, sex, coronary artery disease, heart failure, diabetes, chronic kidney disease, and left ventricular ejection fraction.

Intra-observer variability is the Achilles’ heel of training AI models. When there is significant practice variation in holistic assessment, models trained in one center tend to generalize poorly to others. Moreover, when two clinicians disagree in their interpretation of the same image, the choice of ground-truth label can significantly affect model training (33). Additionally, the choice of threshold used for dichotomization also has a substantial impact on the performance metrics. Within this context, EchoNet-RV was compared to two large multi-task models trained on data from different institutions, both of which provide holistic assessments rather than direct quantitative measurements (20,21). Our results suggest that automated, measurement-driven evaluations continue to hold value, even when large AI models can provide holistic assessments (19). Just as a human cardiologist relies on both quantitative measurements and overall gestalt to arrive at a final clinical decision, AI models that combine measurement-driven and holistic evaluations will be essential for precise assessment of RV function.

EchoNet-RV was designed to mitigate errors arising from beat-to-beat variation by sampling multiple clips from each input video and averaging the individual predictions generated by its spatiotemporal CNN module from these clips to produce the final video-level output. For this test-time augmentation, we evaluated two sampling strategies: one similar to EchoNet-Dynamic’s (13), in which the results of the semantic segmentation guided the sampling of clips, and another, in which 5 clips were sampled randomly from each video. Interestingly, despite its simplicity and computational efficiency, the latter approach achieved marginally superior performance and was therefore adopted in the final EchoNet-RV model. Random sampling ensures that the number of analyzed clips is independent of the number of complete cardiac cycles, provides stable performance even when segmentation is unreliable (e.g., due to poor visibility of the RV free wall), and avoids the propagation of segmentation errors into the downstream task of RVFAC prediction. In addition, it eliminates the need for the segmentation step, thereby reducing processing time and computational demands. Nevertheless, although the semantic segmentation module is not used for RVFAC estimation, we retained it as a key component of EchoNet-RV, as its outputs may be valuable in other applications.

Although several segmentation-free DL models have recently been proposed for predicting RV morphological and functional parameters from echocardiographic videos (20,27,34), EchoNet-RV uniquely enables automated assessment of RVFAC – a key parameter of RV function – and demonstrated consistent performance across a wide range of image qualities, ultrasound machine vendors, and A4C definitions, including both standard and RV-focused views. This robustness underscores the model’s strong generalizability and highlights its potential for seamless integration into diverse real-world clinical environments, regardless of local imaging protocols or equipment.

EchoNet-RV also presents an unparalleled opportunity to enhance the objectivity of RV function assessment, particularly in clinical scenarios where operator bias may inadvertently influence interpretation. In contexts such as long-term disease monitoring or following surgical and transcatheter interventions, clinicians, motivated by the hope for positive outcomes, may unconsciously overestimate functional improvement, especially when faced with ambiguous or technically challenging echocardiographic images. The visual presence of implanted devices or anatomical alterations can also hamper unbiased human evaluation. In contrast, EchoNet-RV offers a consistent, data-driven alternative that is unaffected by clinical expectations. Its ability to provide automated RVFAC measurements ensures standardized RV functional assessment across time points and institutions, enabling more accurate tracking of disease progression and response to therapy. As such, EchoNet-RV can serve as a critical adjunct in both routine clinical care and research. It holds particular promise for integration into point-of-care settings and screening programs, for example, in pulmonary hypertension, and can support an unbiased staging of overall cardiac dysfunction. This, in turn, may improve patient selection for advanced therapies such as left ventricular assist device implantation or transcatheter mitral and tricuspid valve interventions.

### Limitations

Despite the promising results and strengths of our study, several limitations should be acknowledged. First, the inherent uncertainty of human-measured RVFAC values used as the reference standard may have introduced bias, thereby negatively affecting the observed performance, as these measurements represent only an estimate of the true physiological value. Importantly, EchoNet-RV still exhibited robust performance when evaluated using both the original and extended versions of the Bland-Altman analysis that account for uncertainty in the reference values. Moreover, although RVFAC reflects both longitudinal and radial components of RV function, it is unable to capture the ventricle’s anteroposterior motion. Thus, although RVFAC is a guideline-endorsed parameter of RV function with well-established clinical relevance, other metrics that provide a more comprehensive assessment of global RV function and have lower uncertainty may be considered as potential prediction targets in future studies. Second, EchoNet-RV was trained on data originating from a single healthcare system; therefore, the training dataset primarily represents the patient population treated at Stanford Health Care. Nevertheless, EchoNet-RV also demonstrated robust performance in two geographically distinct external datasets, supporting its generalizability and cross-healthcare system reliability. Finally, outcome data were available only for the Semmelweis dataset. Therefore, future studies are warranted to validate the observed associations between EchoNet-RV-predicted RVFAC values and clinical outcomes in larger, multicenter cohorts.

## CONCLUSIONS

EchoNet-RV enables rapid and fully automated segmentation of the RV and assessment of RVFAC from A4C echocardiographic videos, demonstrating robust performance and generalizability across diverse populations. By providing objective, reproducible, and scalable quantification of RV function, EchoNet-RV has strong potential to become a valuable tool for echocardiogram interpretation and longitudinal disease surveillance.

## Data Availability

The datasets analyzed in this study are not publicly available due to privacy and ethical restrictions. The source code and model weights of EchoNet-RV are publicly available at https://github.com/echonet/RV.

## ABBREVIATIONS

A4C: apica 4-chamber view
AI: artificial intelligence
AUC: area under the receiver operating characteristic curve
CNN: convolutional neural network
CV: coefficient of variation
DL: deep learning
ICC: intraclass correlation coefficient
LOA: limit of agreements
MAD: mean absolute difference
MAE: mean absolute error
MDC: minimal detectable change
MMH: MacKay Memorial Hospital
RVFAC: right ventricular fractional area change
TAPSE: tricuspid annular plane systolic excursion
S’: tricuspid annular peak systolic velocity by tissue Doppler imaging
SEM: standard error of measurement

## REFERENCES

1. Hahn RT, Lerakis S, Delgado V et al. Multimodality Imaging of Right Heart Function: JACC Scientific Statement. J Am Coll Cardiol 2023;81:1954–1973.

2. Konstam MA, Kiernan MS, Bernstein D et al. Evaluation and Management of Right-Sided Heart Failure: A Scientific Statement From the American Heart Association. Circulation 2018;137:e578–e622.

3. Schneider M, Aschauer S, Mascherbauer J et al. Echocardiographic assessment of right ventricular function: current clinical practice. Int J Cardiovasc Imaging 2019;35:49–56.

4. Corbett L, O’Driscoll P, Paton M, Oxborough D, Surkova E. Role and application of three-dimensional transthoracic echocardiography in the assessment of left and right ventricular volumes and ejection fraction: a UK nationwide survey. Echo Res Pract 2024;11:8.

5. Soliman-Aboumarie H, Joshi SS, Cameli M et al. EACVI survey on the multi-modality imaging assessment of the right heart European Heart Journal - Cardiovascular Imaging 2022;23:1417–1422.

6. Kovács A, Lakatos B, Tokodi M, Merkely B. Right ventricular mechanical pattern in health and disease: beyond longitudinal shortening. Heart Fail Rev 2019;24:511–520.

7. Kovács A, Magunia H, Nicoara A et al. Challenges and opportunities in assessing right ventricular structure and function: a Roadmap for standardization, clinical implementation and research. Nature Reviews Cardiology 2025.

8. Anavekar NS, Gerson D, Skali H, Kwong RY, Yucel EK, Solomon SD. Two-dimensional assessment of right ventricular function: an echocardiographic-MRI correlative study. Echocardiography 2007;24:452–6.

9. Mukherjee M, Rudski LG, Addetia K et al. Guidelines for the Echocardiographic Assessment of the Right Heart in Adults and Special Considerations in Pulmonary Hypertension: Recommendations from the American Society of Echocardiography. J Am Soc Echocardiogr 2025;38:141–186.

10. Lin LQ, Conway J, Alvarez S et al. Reduced Right Ventricular Fractional Area Change, Strain, and Strain Rate before Bidirectional Cavopulmonary Anastomosis is Associated with Medium-Term Mortality for Children with Hypoplastic Left Heart Syndrome. J Am Soc Echocardiogr 2018;31:831–842.

11. Pinedo M, Villacorta E, Tapia C et al. Inter- and intra-observer variability in the echocardiographic evaluation of right ventricular function. Rev Esp Cardiol 2010;63:802–9.

12. Pavlicek M, Wahl A, Rutz T et al. Right ventricular systolic function assessment: rank of echocardiographic methods vs. cardiac magnetic resonance imaging. Eur J Echocardiogr 2011;12:871–80.

13. Ouyang D, He B, Ghorbani A et al. Video-based AI for beat-to-beat assessment of cardiac function. Nature 2020;580:252–256.

14. Kwan AC, Chang EW, Jain I et al. Deep Learning-Derived Myocardial Strain. JACC Cardiovasc Imaging 2024;17:715–725.

15. Ma J, Yang Z, Kim S, et al. MedSAM2: Segment Anything in 3D Medical Images and Videos. arXiv 2025.

16. He B, Kwan AC, Cho JH et al. Blinded, randomized trial of sonographer versus AI cardiac function assessment. Nature 2023;616:520–524.

17. Tromp J, Bauer D, Claggett BL et al. A formal validation of a deep learning-based automated workflow for the interpretation of the echocardiogram. Nature Communications 2022;13:6776.

18. Duffy G, Cheng PP, Yuan N et al. High-Throughput Precision Phenotyping of Left Ventricular Hypertrophy With Cardiovascular Deep Learning. JAMA Cardiology 2022;7:386–395.

19. Sahashi Y, Ieki H, Yuan V et al. Artificial Intelligence Automation of Echocardiographic Measurements. J Am Coll Cardiol 2025;86:964–978.

20. Holste G, Oikonomou EK, Tokodi M, Kovács A, Wang Z, Khera R. Complete AI-Enabled Echocardiography Interpretation With Multitask Deep Learning. JAMA 2025.

21. Vukadinovic M, Chiu IM, Tang X et al. Comprehensive echocardiogram evaluation with view primed vision language AI. Nature 2025.

22. Chen L-C, Zhu Y, Papandreou G, Schroff F, Adam H. Encoder-Decoder with Atrous Separable Convolution for Semantic Image Segmentation. In: Ferrari V, Hebert M, Sminchisescu C, Weiss Y, editors. Computer Vision – ECCV 2018. Cham: Springer International Publishing, 2018:833–851.

23. Tran D, Wang H, Torresani L, Ray J, LeCun Y, Paluri M. A Closer Look at Spatiotemporal Convolutions for Action Recognition. 2018 IEEE/CVF Conference on Computer Vision and Pattern Recognition 2017:6450–6459.

24. Pasdeloup D, Østvik A, Olaisen S, Skogvoll E, Dalen H, Lovstakken L. Challenges and Strategies for Deep Learning in Cardiovascular Imaging: Ejection Fraction and Heart Failure Management. JACC Cardiovasc Imaging 2025;18:751–764.

25. Kagiyama N, Tokodi M, Hathaway QA et al. PRIME 2.0: Proposed Requirements for Cardiovascular Imaging-Related Multimodal-AI Evaluation: An Updated Checklist. JACC Cardiovasc Imaging 2025.

26. Tokodi M, Szijártó Á. From Promise to Practice: Reducing Research Waste in Deep Learning Model Development for Cardiovascular Imaging. JACC Cardiovasc Imaging 2025;18:765–767.

27. Tokodi M, Magyar B, Soós A et al. Deep Learning-Based Prediction of Right Ventricular Ejection Fraction Using 2D Echocardiograms. JACC Cardiovasc Imaging 2023;16:1005–1018.

28. Myhre PL, Grenne B, Asch FM et al. Artificial intelligence-enhanced echocardiography in cardiovascular disease management. Nat Rev Cardiol 2025.

29. Hsia BC, Lai A, Singh S et al. Validation of American Society of Echocardiography Guideline-Recommended Parameters of Right Ventricular Dysfunction Using Artificial Intelligence Compared With Cardiac Magnetic Resonance Imaging. J Am Soc Echocardiogr 2023;36:967–977.

30. Surkova E, Kovacs A, Tokodi M et al. Contraction Patterns of the Right Ventricle Associated with Different Degrees of Left Ventricular Systolic Dysfunction. Circ Cardiovasc Imaging 2021;14:e012774.

31. Tokodi M, Nemeth E, Lakatos BK et al. Right ventricular mechanical pattern in patients undergoing mitral valve surgery: a predictor of post-operative dysfunction? ESC Heart Fail 2020;7:1246–1256.

32. Bidviene J, Muraru D, Maffessanti F et al. Regional shape, global function and mechanics in right ventricular volume and pressure overload conditions: a three-dimensional echocardiography study. Int J Cardiovasc Imaging 2021;37:1289–1299.

33. Ouyang D. Cardiologists have an AUC of 0.81 with each other. 2023.

34. Szijártó Á, Merkely B, Kovács A, Tokodi M. Deep Learning-Enabled Echocardiographic Assessment of Biventricular Ejection Fractions: The Dual-Task QUEST-EF Model. Eur Heart J Cardiovasc Imaging 2025.

