## Supplementary Appendix for "Artificial Intelligence-Enabled Echocardiographic Assessment of Right Ventricular Function"

**Supplemental Table 1** Clinical characteristics of the Stanford dataset

|  | **All** | **Train** | **Validation** | **Test** |
| --- | --- | --- | --- | --- |
| **Patients**, n | 5,386 | 3,770 | 808 | 808 |
| **Studies**, n | 8,314 | 5,772 | 1,254 | 1,288 |
| **Videos**, n | 8,489 | 5,892 | 1,277 | 1,320 |
| **Age**, years | 59.8  (46.6–71.0) | 59.8  (46.2–71.1) | 59.7  (46.6–71.7) | 59.9  (48.5–70.5) |
| **Males** | 2,745 (51.0) | 1,946 (51.6) | 395 (48.9) | 404 (50.0) |
| **Body mass index**, kg/m^2^ | 26.0  (22.0–30.0) | 26.0  (22.0–30.0) | 25.0  (22.0–31.0) | 25.0  (22.0–30.0) |
| **Body surface area**, m^2^ | 1.82  (1.64–2.01) | 1.82  (1.65–2.01) | 1.81  (1.62–2.00) | 1.82  (1.63–2.01) |
| **Systolic blood pressure**, mmHg | 119 (107–133) | 119 (107–133) | 118 (108–134) | 116 (106–132) |
| **Diastolic blood pressure**, mmHg | 70 (62–78) | 70 (61–79) | 70 (61–78) | 68 (62–77) |
| **Heart rate**, 1/min | 69 (60–102) | 69 (60–102) | 69 (61–102) | 68 (60–101) |
| **Coronary artery disease** | 341 (6.3) | 227 (6.0) | 60 (7.4) | 54 (6.7) |
| **Heart failure** | 1,084 (20.1) | 740 (19.6) | 171 (21.2) | 173 (21.4) |
| **Hypertension** | 1,442 (26.8) | 985 (26.1) | 223 (27.6) | 234 (29.0) |
| **Diabetes** | 708 (13.1) | 483 (12.8) | 119 (14.7) | 106 (13.1) |
| **Chronic kidney disease** | 740 (13.7) | 498 (13.2) | 116 (14.4) | 126 (15.6) |
| **LVEDV**, mL | 82.4  (62.0–109.6) | 83.0  (62.1–109.9) | 79.5  (58.6–106.4) | 82.1  (64.0–110.1) |
| **LVEDVi**, mL/m^2^ | 45.1  (35.0–58.5) | 45.4  (35.1–58.8) | 44.2  (33.7–58.2) | 45.1  (35.5–57.3) |
| **LVESV**, mL | 33.5  (23.6–49.7) | 33.7  (23.6–49.9) | 32.1  (23.1–49.5) | 33.9  (23.9–48.3) |
| **LVESVi**, mL/m^2^ | 18.3  (13.3–26.4) | 18.4  (13.3–26.4) | 17.9  (13.1–26.3) | 18.6  (13.4–26.3) |
| **LVEF**, % | 59.2  (51.2–64.1) | 59.1  (50.8–64.1) | 59.3  (52.1–63.8) | 59.5  (52.3–64.1) |
| **RV end-diastolic area**, cm^2^ | 22.1  (17.4–28.1) | 22.1  (17.5–28.1) | 21.5  (16.9–27.3) | 22.7  (17.6–28.6) |
| **RV end-systolic area**, cm^2^ | 15.3  (11.2–20.8) | 15.3  (11.2–20.8) | 14.8  (10.9–20.6) | 15.6  (11.4–21.1) |
| **RVFAC**, % | 30.1  (21.9–37.4) | 30.0  (22.0–37.4) | 30.3  (21.9–37.4) | 30.3  (21.9–37.8) |

Values are median (interquartile range) or n (%).

LVEDV – left ventricular end-diastolic volume, LVEDVi – left ventricular end-diastolic volume indexed to body surface area, LVEF – left ventricular ejection fraction, LVESV – left ventricular end-systolic volume, LVESVi – left ventricular end-systolic volume indexed to body surface area, RV – right ventricular, RVFAC – right ventricular fractional area change

**Supplemental Table 2** Technical details of the Stanford dataset

|  | **All** | **Train** | **Validation** | **Test** |
| --- | --- | --- | --- | --- |
| **Videos**, n | 8,489 | 5,892 | 1,277 | 1,320 |
| **Frame rate**, 1/sec | 50 (50–51) | 50 (50–51) | 50 (50–52) | 50 (50–52) |
| **Ultrasound machine** |  |  |  |  |
| Philips EPIQ 5G | 5 (0.1) | 1 (0.0) | 3 (0.2) | 1 (0.1) |
| Philips EPIQ 7C | 6,078 (71.6) | 4,196 (71.2) | 922 (72.2) | 960 (72.7) |
| GE Vivid iq | 2 (0.0) | 2 (0.0) | 0 (0.0) | 0 (0.0) |
| Philips iE33 | 2,404 (28.3) | 1,693 (28.7) | 352 (27.6) | 359 (27.2) |

**Supplemental Table 3** Technical details of the videos in the external test sets

|  | **Internal test set** | **Semmelweis** | **MMH** |
| --- | --- | --- | --- |
| **Videos**, n | 1,320 | 3,107 | 1,077 |
| **Frame rate**, 1/sec | 50 (50–52) | 53 (50–60) | 30 (30–48) |
| **Ultrasound machine** |  |  |  |
| GE Vivid 7 | 0 (0.0) | 0 (0.0) | 549 (51.0) |
| GE Vivid E9 | 0 (0.0) | 0 (0.0) | 19 (1.8) |
| GE Vivid E95 | 0 (0.0) | 1,986 (63.9) | 0 (0.0) |
| GE Vivid i | 0 (0.0) | 0 (0.0) | 93 (8.6) |
| Philips EPIQ 5G | 1 (0.1) | 0 (0.0) | 0 (0.0) |
| Philips EPIQ 7C | 960 (72.7) | 358 (11.5) | 25 (2.3) |
| Philips EPIQ 7G | 0 (0.0) | 635 (20.4) | 0 (0.0) |
| Philips EPIQ CVx | 0 (0.0) | 3 (0.1) | 0 (0.0) |
| Philips iE33 | 359 (27.2) | 125 (4.0) | 391 (36.3) |
| **Type of A4C** |  |  |  |
| Standard A4C | - | 1,756 (56.5) | 728 (67.6) |
| RV-focused A4C | - | 1,351 (43.5) | 349 (32.4) |
| **Overall image quality** |  |  |  |
| Excellent | - | 50 (1.6) | 11 (1.0) |
| Good | - | 352 (11.3) | 81 (7.5) |
| Moderate | - | 1,584 (51.0) | 523 (48.6) |
| Poor | - | 1,012 (32.6) | 382 (35.5) |
| Non-diagnostic | - | 109 (3.5) | 80 (7.4) |

Values are median (interquartile range) or n (%).

A4C – apical 4-chamber view, MMH – MacKay Memorial Hospital; other abbreviations as in Supplemental Table 1.

**Supplemental Table 4** Proposed Requirements for Cardiovascular Imaging-Related Multimodal-AI Evaluation (PRIME 2.0) checklist

| **Checklist item** | **Manuscript section/figure/table** |
| --- | --- |
| **1. Designing an AI study in cardiovascular imaging** |  |
| 1.1 Appropriateness of applying AI |  |
| Describe the need for applying AI | Introduction |
| Determine the appropriateness of applying AI | Introduction |
| 1.2 Study objectives, input data type, and prediction target |  |
| Explain the AI task and the likely deployment context | Introduction  Discussion |
| Describe the input data, number of training/test examples | Methods – Datasets  Table 1, Supplemental Tables 1, 2, and 3 |
| Specify model supervision type | Introduction  Methods – Video annotation  Methods – Architecture and development of EchoNet-RV |
| Describe the nature of the model’s output and what it represents | Methods – Video annotation  Methods – Architecture and development of EchoNet-RV |
| 1.3 Design of the AI study |  |
| Describe the study design | Introduction  Central Illustration |
| Describe data origin | Methods – Datasets |
| Describe whether impact analysis was included | Impact analysis was not performed. |
| **2. Data format and preprocessing** |  |
| 2.1 Data format |  |
| Describe the technical details of the data acquisition | Methods – Datasets  Methods – Video annotation  Supplemental Tables 2 and 3 |
| Describe the technical details of the data format | Methods – Video preprocessing |
| 2.2 Clinical characteristics of the study cohort |  |
| Present the age, sex, and race distributions of the cohorts | Table 1, Supplemental Table 1 |
| Summarize key clinical and imaging characteristics of the cohorts | Table 1, Supplemental Tables 1, 2, and 3 |
| Compare summary statistics of cases and controls | N/A |
| 2.3 Steps of data preprocessing |  |
| Describe how data were cleaned, made uniform, and consistent | Methods – Dataset  Methods – Video annotation  Methods – Video preprocessing |
| Describe data harmonization techniques (if applicable) | N/A |
| Provide details on missing values and imputation methods | N/A |
| Describe processes for handling outliers | Methods – Video preprocessing |
| Describe whether class imbalance exists | Results – Performance in identifying RV dysfunction |
| **Checklist item** | **Manuscript section/figure/table** |
| 2.4 Feature engineering and feature selection |  |
| Describe applied feature engineering techniques | N/A |
| Describe applied feature selection techniques | N/A |
| **3. Selection of AI methods and applications** |  |
| 3.1 Selecting appropriate AI methods and applications |  |
| Clearly define data composition (structured/unstructured) | Methods – Datasets  Methods – Video annotation |
| 3.2–3.4 Training strategies |  |
| Describe the AI method used, with rationale for clinical task fit | Methods – Architecture and development of EchoNet-RV |
| 3.5 Solving clinical problems | Introduction  Methods – Architecture and development of EchoNet-RV |
| **4. Model assessment** |  |
| 4.1 Importance of model assessment |  |
| Describe the evaluation approach and how it addresses the clinical question | Methods – Performance metrics and statistical analysis |
| 4.2 Technical performance metrics |  |
| Report relevant performance metrics | Results (entire section)  Tables 2, 3, and 4  Figures 1, 3, 4, and 5  Supplemental Tables 5 and 6 |
| Justify metric selection based on task characteristics | Methods – Performance metrics and statistical analysis |
| Describe manual annotation process, reference-standard prep, and observer variability | Methods – Video annotation  Methods – Comparing prediction error with beat-to-beat and inter-observer variability  Results – Performance in RVFAC prediction  Tables 2 and 3 |
| 4.3–4.4 Robustness, generalizability, and evaluating data quality |  |
| Evaluate model robustness to external variations | Results (entire section)  Tables 2, 3, and 4  Figures 1, 3, 4, and 5  Supplemental tables 5 and 6 |
| Assess performance across clinical subpopulations and sociodemographic subgroups |  |
| 4.5 Identifying features learned by the model |  |
| Describe interpretability/explainability methods | Semantic segmentation is inherently explainable.  Explainability of the spatiotemporal CNN model was not explored. |
| Quantify and discuss uncertainty | Discussion – Limitations |

| **Checklist item** | **Manuscript section/figure/table** |
| --- | --- |
| **5. Clinical evaluation** |  |
| 5.1 Importance of clinical evaluation |  |
| Describe potential clinical impact of misclassification | Misclassification could lead to delayed diagnosis, inappropriate risk stratification, and suboptimal patient management. |
| 5.2 Clinical utility metrics |  |
| Define clinical utility of the AI system | Results (entire section)  Tables 2, 3, 4, 5, and 6  Central Illustration  Figures 1, 3, 4, 5, and 6  Supplemental Tables 5 and 6  Discussion |
| Describe cost-effectiveness analysis | Cost-effectiveness analysis was not performed. |
| 5.3 Clinical validation |  |
| Provide evidence of clinical validation | Results (entire section)  Tables 2, 3, 4, 5, and 6  Figures 1, 3, 4, 5, and 6  Supplemental Tables 5 and 6 |
| 5.4 Continuous monitoring |  |
| Outline plans for post-deployment monitoring | EchoNet-RV’s performance will be assessed in additional validation cohorts.  If performance degradation is detected, the precise cause will be identified and the model will be refined (e.g., by updating training data [retraining], model parameter values [recalibrating], or improving the modeling structure or training method). |
| **6. Best practices for replicability** |  |
| 6.1 Importance of transparency and open science principles |  |
| Ensure data sharing follows FAIR (findability, accessibility, interoperability, and reusability) principles | Data could not be shared due to proprietary and institutional restrictions. |
| Report on training data representativeness | Methods – Datasets  Table 1  Supplemental Table 1 |
| 6.2 Ensuring technical reproducibility |  |
| Report results for the final model and training process | Methods – Architecture and development of EchoNet-RV  Results (entire section)  Tables 2, 3, and 4  Figures 1, 3, 4, and 5  Supplemental Tables 5 and 6 |
| Describe sources of randomness in training |  |
| Report random seeds (if applicable) |  |
| Describe hardware setup |  |
| Evaluate uncertainty from randomness |  |
| Justify if source code, model weights, or datasets are not shared | The source code and model weights are publicly available:  <https://github.com/echonet/RV> |
| 6.3 Specific considerations for reproducing large language models and generative AI studies | N/A |

| **Checklist item** | **Manuscript section/figure/table** |
| --- | --- |
| **7. Reporting of limitations, biases, and alternatives** |  |
| 7.1 Acknowledging study and model limitations |  |
| Discuss key limitations (data, methodology, generalizability) | Discussion – Limitations |
| Report sensitivity analyses and model-specific issues | Results (entire section)  Tables 2, 3, 4, 5, and 6  Figures 1, 3, 4, 5, and 6  Supplemental Tables 5 and 6 |
| 7.2 Discussing study strengths |  |
| Articulate strengths (methodological rigor, dataset quality, innovation, clinical relevance) | Discussion |
| 7.3 Reporting on bias and fairness assessment |  |
| Report stratified performance metrics for demographic subgroups | Results (entire section)  Tables 2, 3, 4, 5, and 6  Figures 1, 3, 4, 5, and 6  Supplemental Tables 5 and 6 |
| Describe bias mitigation strategies and residual bias | Discussion – Limitations |
| Acknowledge fairness evaluation limitations | Discussion – Limitations |
| 7.4 Contextualizing with alternative methods |  |
| Benchmark against clinical standards/traditional risk scores | Methods – Architecture and development of EchoNet-RV  Methods – Benchmarking against multi-task models  Results – Performance in RVFAC prediction  Results – Benchmarking against multi-task models  Table 4 |
| Justify complex AI models over simpler alternatives |  |
| Discuss complementary methods for validation | N/A |

AI – artificial intelligence, CNN – convolutional neural network, DL – deep learning; other abbreviations as in Supplemental Table 1.

**Supplemental Table 5** Performance of EchoNet-RV in different subgroups of the external test sets

|  | **Semmelweis** | | | | **MMH** | | | |
| --- | --- | --- | --- | --- | --- | --- | --- | --- |
|  | **N of videos** | **MAE** | **ICC** | **AUC** | **N of videos** | **MAE** | **ICC** | **AUC** |
| **All** | 3,107 | 5.830  (5.670–5.992) | 0.481  (0.452–0.509) | 0.727  (0.709–0.744) | 1,077 | 6.362  (6.064–6.660) | 0.301  (0.243–0.356) | 0.684  (0.648–0.719) |
| **Ultrasound system** |  |  |  |  |  |  |  |  |
| GE | 1,986 | 5.381  (5.194–5.573) | 0.724  (0.702–0.746) | 0.487  (0.450–0.521) | 661 | 6.100  (5.736–6.468) | 0.256  (0.184–0.324) | 0.661  (0.611–0.709) |
| Philips | 1,121 | 6.626  (6.330–6.924) | 0.726  (0.696–0.756) | 0.462  (0.416–0.506) | 416 | 6.778  (6.284–7.294) | 0.317  (0.229–0.400) | 0.689  (0.634–0.743) |
| **Type of A4C view** |  |  |  |  |  |  |  |  |
| Standard | 1,756 | 5.721  (5.513–5.935) | 0.495  (0.457–0.531) | 0.719  (0.695–0.743) | 728 | 6.649  (6.267–7.030) | 0.212  (0.139–0.282) | 0.650  (0.603–0.696) |
| RV-focused | 1,351 | 5.973  (5.714–6.226) | 0.460  (0.416–0.503) | 0.744  (0.718–0.770) | 349 | 5.764  (5.318–6.198) | 0.471  (0.388–0.544) | 0.762  (0.705–0.816) |
| **Overall image quality** |  |  |  |  |  |  |  |  |
| Excellent | 50 | 4.846  (3.792–5.948) | 0.550  (0.327–0.707) | 0.686  (0.526–0.833) | 11 | 5.337  (2.953–8.119) | 0.893  (-) | 0.551  (-0.044 to 0.868) |
| Good | 352 | 5.093  (4.705–5.500) | 0.562  (0.484–0.629) | 0.756  (0.705–0.803) | 81 | 6.250  (5.251–7.299) | 0.720  (0.588–0.839) | 0.418  (0.241–0.561) |
| Moderate | 1,584 | 5.682  (5.459–5.906) | 0.492  (0.451–0.531) | 0.721  (0.696–0.746) | 523 | 6.167  (5.767–6.577) | 0.712  (0.659–0.765) | 0.326  (0.251–0.396) |
| Poor | 1,012 | 6.341  (6.036–6.658) | 0.447  (0.397–0.494) | 0.730  (0.699–0.760) | 382 | 6.548  (6.036–7.080) | 0.629  (0.565–0.692) | 0.235  (0.123–0.336) |
| Non-diagnostic | 109 | 6.083  (5.213–7.010) | 0.404  (0.241–0.542) | 0.696  (0.594–0.795) | 80 | 7.004  (5.751–8.323) | 0.577  (0.448–0.704) | 0.057  (-0.150 to 0.248) |

Performance metrics are reported with their corresponding 95% confidence intervals computed using 10,000 bootstrapped samples. MAEs are expressed as percentage points. AUCs are reported for identifying videos with an actual RVFAC of less than 35%.

AUC – area under the receiver operating characteristic curve, ICC – intra-class correlation coefficient, MAE – mean absolute error; other abbreviations as in Supplemental Tables 1 and 3.

**Supplemental Table 6** Performance of EchoNet-RV in identifying RV dysfunction

|  | **AUC** | **Accuracy** | **Specificity** | **Sensitivity** | **NPV** | **PPV** |
| --- | --- | --- | --- | --- | --- | --- |
| **Internal test set** | 0.859  (0.843–0.876) | 0.792  (0.770–0.813) | 0.645  (0.600–0.690) | 0.866  (0.843–0.887) | 0.707  (0.671–0.745) | 0.829  (0.811–0.847) |
| **Semmelweis** (video-level) | 0.725  (0.710–0.740) | 0.653  (0.637–0.669) | 0.719  (0.695–0.743) | 0.600  (0.577–0.623) | 0.588  (0.573–0.605) | 0.729  (0.711–0.748) |
| **Semmelweis** (study-level) | 0.770  (0.746–0.794) | 0.699  (0.670–0.726) | 0.769  (0.730–0.807) | 0.644  (0.602–0.684) | 0.626  (0.598–0.656) | 0.782  (0.751–0.813) |
| **MMH** | 0.684  (0.653–0.713) | 0.672  (0.644–0.699) | 0.727  (0.696–0.757) | 0.513  (0.455–0.571) | 0.813  (0.794–0.832) | 0.392  (0.354–0.430) |

Performance metrics are reported with their corresponding 95% confidence intervals computed using 10,000 bootstrapped samples. AUCs are reported for identifying videos/studies with an actual RVFAC of less than 35%. Accuracy, specificity, sensitivity, negative predictive value, and positive predictive value were computed after dichotomizing the predicted and actual RVFAC values using the guideline-recommended cutoff value of 35%.

NPV – negative predictive value, PPV – positive predictive value; other abbreviations as in Supplemental Tables 1, 3, and 5.
